# Leveraging Electronic Health Records to Investigate Sex Differences in Oral Diseases and Conditions

**DOI:** 10.1101/2024.11.10.24317064

**Authors:** E. Fetchko, L. Sangalli, A. Letra

## Abstract

**Objectives:** Sexual dimorphism has been shown to influence disease predisposition and/or progression, however, studies addressing sex-based differences in dental, oral, and craniofacial (DOC) diseases and conditions are scarce. This study aimed to identify DOC diseases and conditions likely influenced by sexual dimorphism using two large data repositories.

**Methods:** Retrospective study of medical/dental record data obtained from adult participants (>18 years old) in the NIH *All of Us* Research Program (n=254,700) and the BigMouth Data Repository (n ≈ 4.7 million). The number of males and females presenting each selected DOC disease concept in each database was recorded. Sex-specific association analysis for each concept was performed using chi-square tests (α ≦ 0.0002). Female-to-male odds ratio (OR) and confidence intervals were also calculated.

**Results:** The initial search in *All of Us* and BigMouth yielded 216 and 243 DOC concepts, respectively. Eighty-seven of 216 dental concepts identified in *All of Us* had sex-stratified data. Of these, significant sex-related differences were found for 61/87 concepts (70%), with 33 concepts (54%) showing female bias and 28 (46%) showing male bias (P≦ 0.0002). Higher female bias was noted for ‘diseases of oral soft tissues’, ‘disorders of tooth development and eruption’, and ‘diseases of pulpal/periapical tissues’, whereas higher male bias was noted for ‘gingival and periodontal diseases’, ‘dental caries’, and ‘malignant tumor of oral cavity’(P≦ 0.0002). Analysis of BigMouth data showed sex bias for 90/230 (39%) concepts investigated, of which 87 (97%) showed female bias and 3 (3%) showed male bias (P≦ 0.0002). Discordant sex bias results among the databases were noted for 8 concepts.

**Conclusions:** This study provides evidence of sex bias in numerous DOC diseases and conditions in the populations studied. Additional studies in other populations and considering sociodemographic factors might provide further insight into the role of sexual dimorphism in DOC diseases.

## Introduction

Sex is a biological variable that accounts for differences between males and females across multiple species and cellular domains such as growth, metabolism, reproduction, and aging processes (Manandhar et al. 2018). These differences may confer increased vulnerability for specific infections, diseases, and conditions in one sex over the other (Ubeda and Janse 2016). Previous studies have found significant differences in immune functions between males and females that influence the course and manifestation of infections (Klein 2000). Males typically exhibit a more robust acute inflammatory response, while females display attenuated inflammatory reactions (Valerio and Kirkwood 2018). This divergence is a consistent trait among mammals, where the innate immune response significantly differs between sexes (Valerio and Kirkwood 2018). Moreover, studies have indicated that sex hormones exert differential effects on the immune system, contributing to distinct risk profiles for certain disorders in males and females (Keselman and Heller 2015). In addition, specific genes on the X chromosome are shown to influence metabolism, injury response, behavior (Arnold et al. 2015; Keogh and Boerner 2020; Lipsky et al. 2021), as well as expression of cytokines, and other immune response mediators (Klein and Flanagan 2016). More recently, a growing body of research has highlighted significant differences in disease manifestations between males and females, driven by these underlying biological mechanisms (Mauvais-Jarvis 2020).

Yet, large clinical studies or studies utilizing human data examining sex-related implications in oral, dental, and craniofacial diseases and disorders are scarce. Although oral diseases affect almost half of the population worldwide (World Health Organization 2022), the understanding of sex-specific manifestations that could confer differential risk for certain oral conditions remains limited. These variations in oral manifestations could be attributed to differences in immune function, hormonal influences, or genetic predisposition (Sangalli et al. 2023). Without a clear understanding of which conditions show a different manifestation in males and females, grasping the underlying cause becomes challenging.

Numerous studies have shown contrasting biological traits between men and women, including certain investigations into specific oral conditions and sex-related distinctions. Some examples include apical periodontitis and dental caries. Studies of apical periodontitis have yielded mixed outcomes, with findings supporting no significant difference between the sexes, while others suggesting a higher prevalence in women (Al-Naxhan et al. 2017; Berlink et al. 2015). Similarly, studies focused on dental caries have conflicting findings, with some indicating female predilection (Lukacs and Largaespada 2006; Lukacs 2011), others favoring males (Furuta et al. 2010), and others failing to demonstrate any sex-related differences (Shaffer et al. 2015). Discrepancies among these results are likely due to differences in sample sizes, parameters evaluated, and populations studied. Therefore, additional studies with large sample sizes are warranted to determine the oral and dental conditions likely affected by sexual dimorphism.

The National Institute of Health (NIH) *All of Us* Research Program (*All of Us*) (The *All of Us* Research Program Investigators, 2019) consists of a national effort to collect medical, dental, and sociodemographic data, as well as biological samples from one million people in the United States to foster advancements in precision medicine and improve health. Through the *All of Us* Researcher Workbench, investigators can access electronic health record (EHR), survey, and genomic data of program participants in completely de-identified format to conduct research. It constitutes a valuable publicly available tool for conducting studies leveraging a large sample size aimed at enhancing personalized medicine approaches (Ramirez et al. 2022). Researchers can query the database for specific disease/condition concepts and obtain data on the total number of participants presenting with each concept, their male-female distributions, race/ethnicity, and age ranges. Researchers may be approved to use *All of Us* data after signing a data use agreement and having completed research ethics training modules. Similarly, the BigMouth Dental Repository (BigMouth) is a centralized dental data repository derived from a national consortia of US dental schools (Consortium of Oral Health Research and Informatics, COHRI) (Walji et al. 2014). Launched in 2012, BigMouth contains de-identified EHR data obtained from dental diagnostic and procedure codes, as well as self-reported medical history data from patients seen at each of the current 11 participating dental schools. BigMouth is not publicly available; access is provided to those institutions that contribute to the repository. While *All of Us* and BigMouth have inherent differences due to their sample population and types of data available, both databases offer immense potential for research in dental, oral, and craniofacial (DOC) research and evidence-based dentistry on a large scale.

In this study, we leveraged the availability of large-scale data from *All of Us* and BigMouth to investigate common DOC diseases and conditions likely influenced by sexual dimorphism. The results highlight numerous conditions for which sexual dimorphism may play a role in the pathogenesis and provide insight into the development of future precision-based diagnostic and treatment strategies.

## Materials and Methods

### Data sources

This retrospective study used medical/dental record data from adult participants (aged 18-89 years old) obtained via the NIH *All of Us* Research Program (n= 254,700) and the BigMouth Dental Data Repository (n≈ 4.7 million). Data search was conducted by a single investigator (EF) between May and August, 2023.

A comprehensive set of search terms was built around common DOC diseases/conditions. For *All of Us*, search terms were informed by the Systemized Nomenclature of Medicine (SNOMED, https://www.snomed.org/) identification number or International Classification of Diseases (ICD)-10 (https://www.cdc.gov/nchs/icd/icd10.htm), and included “oral carcinoma”, “oral cancer”, “tongue cancer”, “oral tumor”, “periodontitis”, “periodontal disease”, “dental”, “dental disease”, “tooth”, “oral”, “mouth”, “infection”, “caries”, “dental anomaly”. The same search terms were utilized for the BigMouth queries to identify diagnostic codes associated with each concept.

The participant count for each DOC concept in each database was then recorded and stratified by biological sex (male, female, or other). By default, exact participant counts for EHR concepts with fewer than 20 participants are not provided in *All of Us to* protect participant privacy, therefore comparisons in such cases were not performed. Similarly, concepts showing fewer than 10 participants were not included in the analysis. The study workflow is presented in Figure 1.

**Figure 1.**
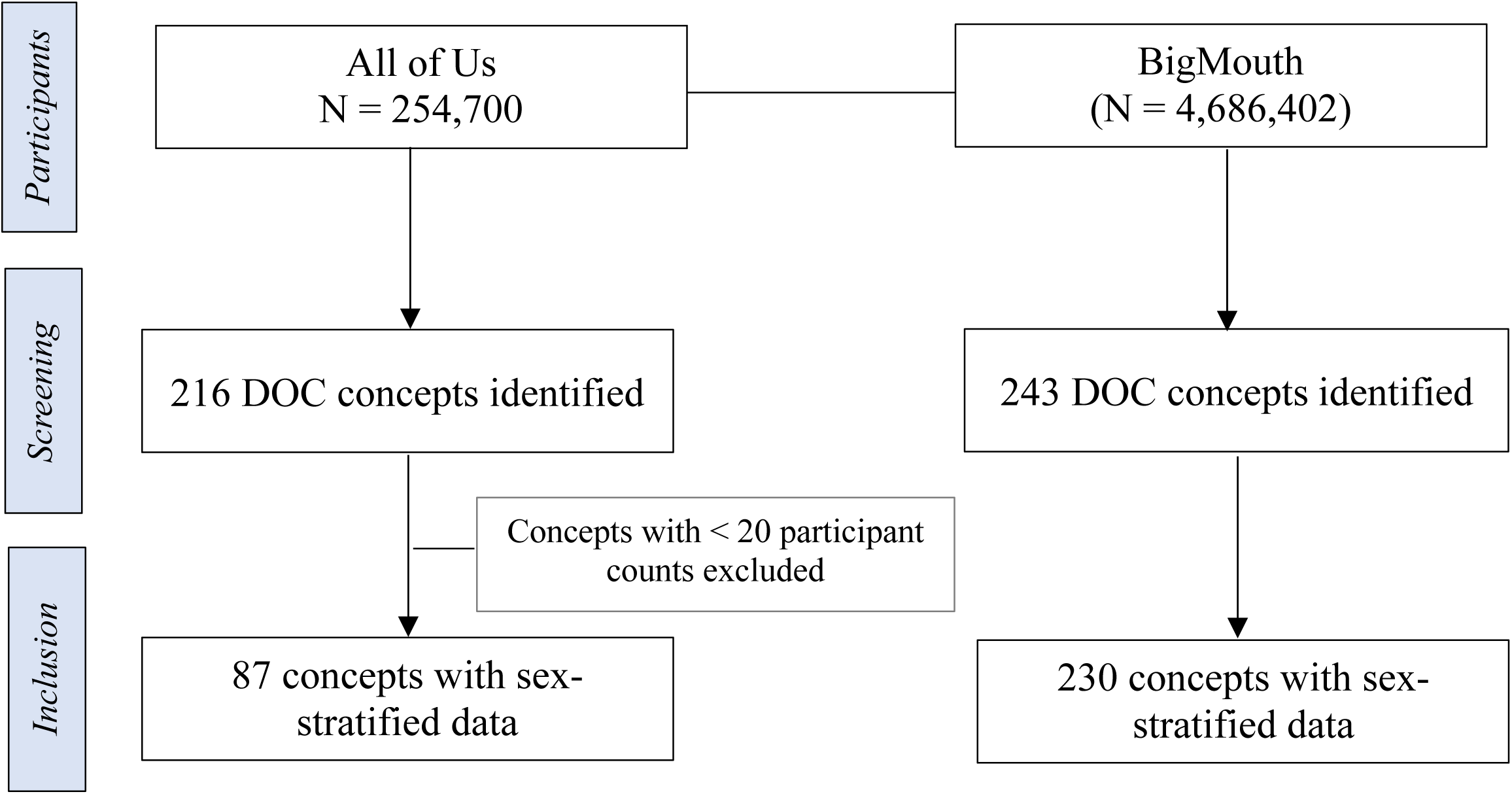
Study workflow for inclusion of dental, oral, and craniofacial concepts investigated.

### Statistical analysis

Sex-specific association analysis for each DOC disease concept was performed using chi-square tests, with a Bonferroni-corrected p≦ 0.0002 denoting statistical differences. Female-to-male odds ratio (OR) and 95% confidence intervals (95% CI) were also calculated to compare the relative odds of the occurrence of each concept in females and males. All statistical analyses were performed using the free online tool SISA (Simple Interactive Statistical Analysis, https://www.quantitativeskills.com/sisa/).

## Results

### Sex differences in DOC concepts from All of Us

A total of 216 DOC concepts were identified in *All of Us*, of which 87 had sex-stratified frequency data. Of these, significant sex-related differences were found for 61/87 concepts (70%), with 33 concepts (54%) showing female bias and 28 (46%) showing male bias (p≦0.0002). The top 3 concepts showing significant statistical evidence of female bias were ‘diseases of the oral soft tissues’ [recurrent aphtous stomatitis (p≦ 0.0001; OR = 5.37), aphtous ulcer of mouth (p≦ 0.0001; OR = 5.36), xerostomia (p≦ 0.0001; OR = 4.96)], followed by ‘disorders of tooth development and eruption’ [anodontia (p≦ 0.0001; OR = 5.29), disturbance of tooth eruption or exfoliation (p≦ 0.0001; OR= 5.14), dental arch relationship (p≦ 0.0001; OR = 4.00)], and ‘diseases of pulp and periapical tissues’ [dental abscess (p≦ 0.0001; OR= 2.78), reversible pulpitis (p≦ 0.0001; OR=2.68), toothache p≦ 0.0001; OR=2.56)]. For ‘gingival and periodontal diseases’, aggressive periodontitis showed the strongest statistical evidence of female bias (p≦ 0.0001; OR=2.56), followed by chronic periodontitis (p≦ 0.0001; OR=1.33), and accretion of teeth (p≦ 0.0001; OR=1.29). ‘Dental caries’ and ‘dental caries on smooth surface penetrating into dentin’ also showed statistical evidence of female bias (p≦ 0.0001; OR=1.33 and OR=1.29, respectively). Conditions in the ‘others’ category showing significant female bias were unsatisfactory tooth restoration (p≦ 0.0001; OR=4.00), and alveolitis of jaw (p≦ 0.0001; OR=3.45) (Table 1).

**Table 1.**
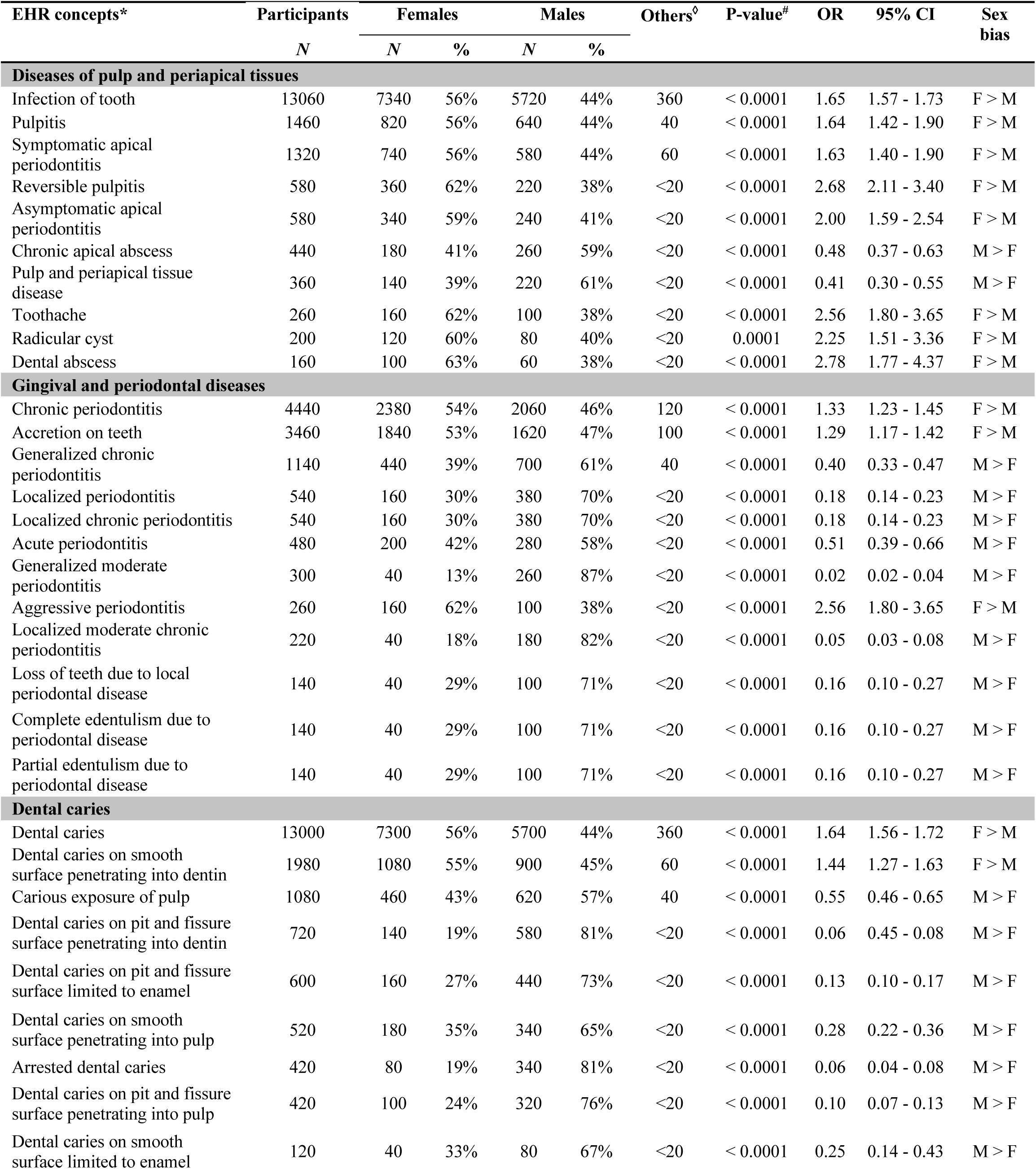
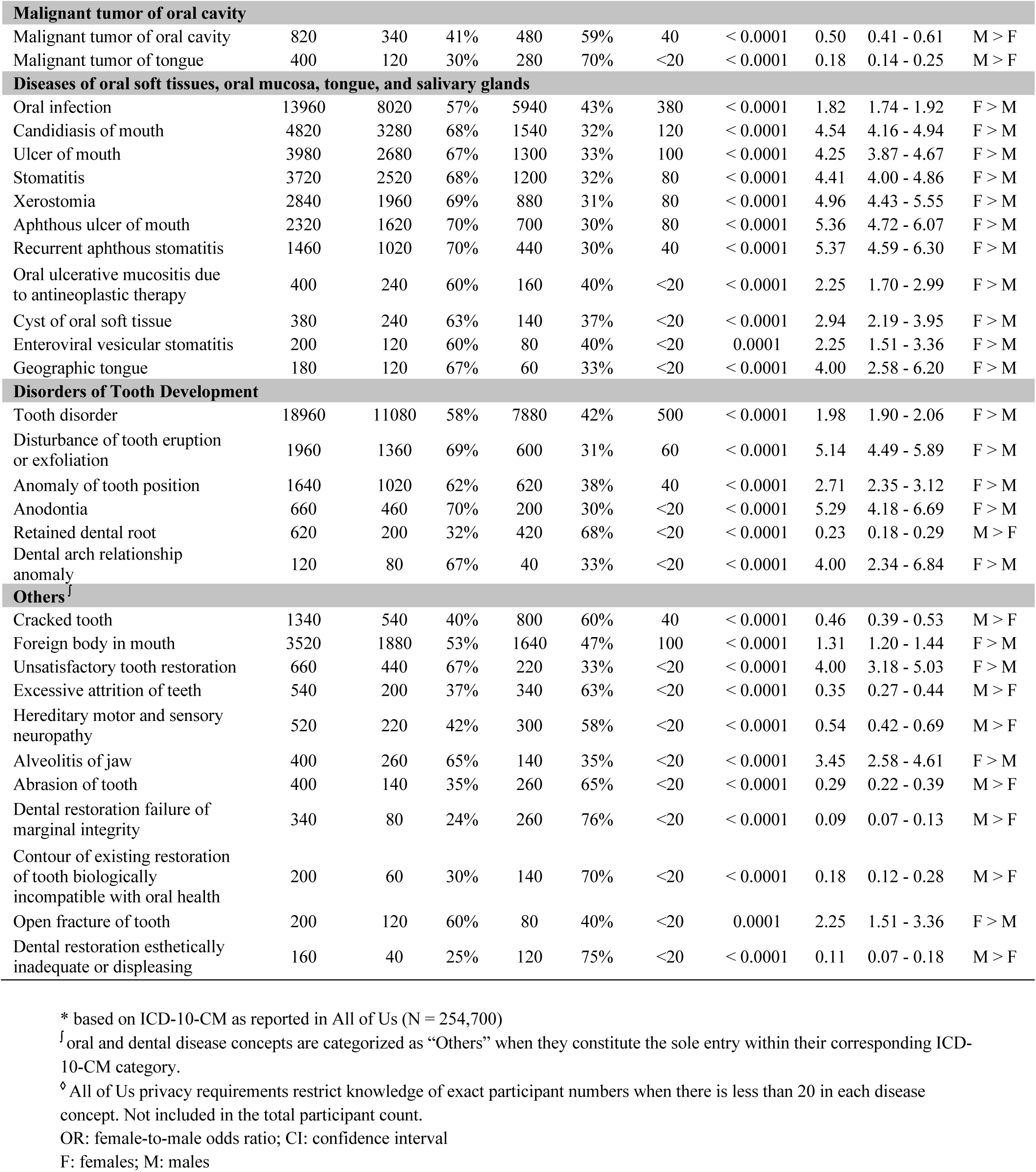
Dental, oral and craniofacial (DOC) concepts extracted from *All of Us* showing statistically significant sex differences.

In contrast, the 28 concepts exhibiting a male bias reflect a majority of concepts in ‘gingival and periodontal diseases’ [generalized moderate periodontitis (p≦ 0.0001; female to male OR=0.02), localized moderate periodontitis (p≦ 0.0001; OR=0.05), and tooth loss due to local periodontal disease (p≦ 0.0001; female to male OR=0.16)], followed by ‘dental caries’ concepts [dental caries on pit and fissure penetrating into dentin (p≦ 0.0001; female to male OR=0.06), dental caries on pit and fissure penetrating into pulp (p≦ 0.0001; female to male OR=0.10)]. Male bias was also observed for ‘malignant tumor of oral cavity’ concepts (p≦ 0.0001; female to male OR=0.18). (Table 1).

Additional concepts showing significant sex bias in All of Us data are shown in Table 1. Concepts in which a trend for significance (0.0002 ≦ p ≦ 0.05) for sex bias or non-significant results were observed are shown in Appendix Table 1.

### Sex differences in DOC concepts from BigMouth

We used BigMouth to validate findings obtained via *All of Us*. Using the same search criteria, we identified 243 oral and dental diseases concepts in BigMouth that were mostly related to the concepts identified in *All of Us*. Of these, 230 concepts had sex-specific data, with 90/230 (39%) showing statistically significant differences among the sexes (p ≦ 0.0002); 87 (97%) showing female bias and 3 (3%) showing male bias (p≦0.0002) (Table 2). The top 3 concepts showing a predominantly female bias were ‘disorders of tooth development and eruption’ [cementum defect (p≦ 0.0001; OR = 5.39), dentin defect (p≦ 0.0001; OR= 3.37), enamel defect (p≦ 0.0001; OR = 2.56)], ‘diseases of the oral soft tissues, oral mucosa, tongue, and salivary glands’[xerostomia (p≦ 0.0001; OR = 5.93), red-blue lesion (p≦ 0.0001; OR = 4.47), tongue swelling/pathology (p≦ 0.0001; OR = 3.61)], and ‘diseases of pulp and periapical tissues’ [symptomatic apical periodontitis (p≦ 0.0001; OR=2.77), symptomatic reversible pulpitis (p≦ 0.0001; OR=2.64), and previously treated teeth (p≦ 0.0001; OR = 2.45)]. For ‘disorders of teeth, hard tissues of teeth, and supporting structures’, significant female bias was observed for temporomandibular disorders (p≦ 0.0001; OR=9.72), sensitive dentin (p≦ 0.0001; OR=3.40), and primary occlusal trauma (p≦ 0.0001; OR=3.31). Other conditions predominantly showing female bias were harmful oral habits (p≦ 0.0001; OR = 2.34), systemic disease manifestation in jaw (p≦ 0.0001; OR = 2.19)], and benign odontogenic pathology (p≦ 0.0001; OR = 2.10)] (Table 2).

**Table 2.** Dental, oral and craniofacial concepts in BigMouth showing sex bias.

| EHR concept* | Participants | Females |  | Males |  | Others<sup>o</sup> | P value<sup>#</sup> | OR | 95% CI | Sex bias |
| --- | --- | --- | --- | --- | --- | --- | --- | --- | --- | --- |
|  | N | N | % | N | % |  |  |  |  |  |
| Diseases of pulp and periapical tissues |  |  |  |  |  |  |  |  |  |  |
| Periapical diagnosis | 20310 | 11818 | 58% | 8284 | 41% | 208 | <0.0001 | 2.02 | 1.94 - 2.10 | F > M |
| Pulp necrosis | 19296 | 10288 | 53% | 8802 | 46% | 206 | <0.0001 | 1.36 | 1.31 - 1.42 | F > M |
| Previously endodontically treated | 14414 | 8734 | 61% | 5553 | 39% | 127 | <0.0001 | 2.45 | 2.34 - 2.57 | F > M |
| Symptomatic apical periodontitis | 9818 | 6072 | 62% | 3626 | 37% | 120 | <0.0001 | 2.77 | 2.61- 2.93 | F > M |
| Asymptomatic apical periodontitis | 4294 | 2403 | 56% | 1858 | 43% | 33 | <0.0001 | 1.67 | 1.53 - 1.81 | F > M |
| Asymptomatic irreversible pulpitis | 3789 | 2077 | 55% | 1678 | 44% | 34 | <0.0001 | 1.53 | 1.39 - 1.67 | F > M |
| Normal apical tissues | 3791 | 2187 | 58% | 1571 | 41% | 33 | <0.0001 | 1.93 | 1.76 – 2.11 | F > M |
| Chronic apical abscess | 3398 | 1816 | 53% | 1556 | 46% | 26 | <0.0001 | 1.36 | 1.24 - 1.49 | F > M |
| Previously initiated therapy | 2949 | 1775 | 60% | 1144 | 39% | 30 | <0.0001 | 2.39 | 2.15 - 2.65 | F > M |
| Normal pulp | 2589 | 1440 | 56% | 1118 | 43% | 31 | <0.0001 | 1.65 | 1.48 - 1.84 | F > M |
| Acute apical abscess | 2379 | 1271 | 53% | 1092 | 46% | 16 | <0.0001 | 1.35 | 1.21 - 1.52 | F > M |
| Symptomatic reversible pulpitis | 1699 | 1044 | 61% | 640 | 38% | 15 | <0.0001 | 2.64 | 2.30 - 3.03 | F > M |
| Asymptomatic reversible pulpitis | 956 | 528 | 55% | 418 | 44% | 10 | <0.0001 | 1.59 | 1.33 - 1.90 | F > M |
| Gingival and periodontal diseases |  |  |  |  |  |  |  |  |  |  |
| Chronic periodontitis | 127902 | 63629 | 50% | 60270 | 47% | 4003 | <0.0001 | 1.11 | 1.09 - 1.13 | F > M |
| Generalized slight chronic periodontitis | 59390 | 28678 | 48% | 27080 | 46% | 3632 | <0.0001 | 1.11 | 1.09 - 1.14 | F > M |
| Plaque induced gingival disease without local contributing factors | 38113 | 21295 | 56% | 16587 | 44% | 231 | <0.0001 | 1.64 | 1.60 - 1.69 | F > M |
| Generalized moderate chronic periodontitis | 31787 | 16251 | 51% | 15387 | 48% | 149 | <0.0001 | 1.12 | 1.08 - 1.15 | F > M |
| Periodontal health | 30215 | 16967 | 56% | 13183 | 44% | 65 | <0.0001 | 1.65 | 1.60 - 1.71 | F > M |
| Generalized severe chronic periodontitis | 22199 | 10248 | 46% | 11838 | 53% | 113 | <0.0001 | 0.75 | 0.72 - 0.78 | M > F |
| Plaque induced gingival disease with local contributing factors | 20461 | 11304 | 55% | 9091 | 44% | 66 | <0.0001 | 1.54 | 1.49 - 1.61 | F > M |
| Localized moderate chronic periodontitis | 19096 | 10343 | 54% | 8678 | 45% | 75 | <0.0001 | 1.42 | 1.36 - 1.48 | F > M |
| Localized severe chronic periodontitis | 15029 | 7649 | 51% | 7326 | 49% | 54 | 0.0002 | 1.09 | 1.04 - 1.14 | F > M |
| Healthy periodontium with attachment loss | 14764 | 7695 | 52% | 7042 | 48% | 27 | <0.0001 | 1.19 | 1.14 - 1.25 | F > M |
| Localized slight chronic periodontitis | 6204 | 3502 | 56% | 2684 | 43% | 18 | <0.0001 | 1.20 | 1.14 - 1.25 | F > M |
| Gingival soft tissue recession-facial and lingual surfaces | 1594 | 946 | 59% | 647 | 41% | 1 | <0.0001 | 2.14 | 1.86 - 2.46 | F > M |
| Gingival diagnosis non-plaque induced | 1489 | 926 | 62% | 563 | 38% | 0 | <0.0001 | 2.71 | 2.33 - 3.14 | F > M |
| Mucogingival deformity- lack of keratinized gingiva | 813 | 491 | 60% | 322 | 40% | 0 | <0.0001 | 2.33 | 1.91 - 2.84 | F > M |
| Gingival swelling | 787 | 473 | 60% | 310 | 39% | 4 | <0.0001 | 2.32 | 1.89 - 2.84 | F > M |
| Insufficient biological width | 626 | 402 | 64% | 224 | 36% | 0 | <0.0001 | 3.22 | 2.56 - 4.06 | F > M |
| Inflammatory jaw lesion | 536 | 306 | 57% | 224 | 42% | 6 | <0.0001 | 1.85 | 1.45 - 2.36 | F > M |
| Gingivitis associated with lichen planus | 449 | 349 | 78% | 100 | 22% | 0 | <0.0001 | 12.18 | 8.89 -16.68 | F > M |
| Lack of gingiva/keratinized tissue | 425 | 267 | 63% | 158 | 37% | 0 | <0.0001 | 2.86 | 2.16 - 3.77 | F > M |
| Localized slight aggressive periodontitis | 192 | 114 | 59% | 76 | 40% | 2 | 0.0001 | 2.23 | 1.48 - 3.36 | F > M |
| Gingival hyperplasia | 166 | 103 | 62% | 62 | 37% | 1 | <0.0001 | 2.74 | 1.76 - 4.28 | F > M |
| Gingivitis associated with pemphigoid | 119 | 85 | 71% | 34 | 29% | 0 | <0.0001 | 6.25 | 3.56 -10.97 | F > M |
| Other soft tissue enlargement | 109 | 72 | 66% | 36 | 33% | 1 | 0.000001 | 3.95 | 2.25 - 6.93 | F > M |
| Gingivitis of genetic origin | 109 | 69 | 63% | 40 | 37% | 0 | 0.0001 | 2.98 | 1.72 - 5.16 | F > M |
| Gingivitis associated with mucocutaneous disorder | 106 | 75 | 71% | 31 | 29% | 0 | <0.0001 | 5.85 | 3.24 -10.58 | F > M |
| Pyogenic granuloma of gingiva | 82 | 53 | 65% | 28 | 34% | 1 | 0.0001 | 3.53 | 1.85 - 6.70 | F > M |
| Gingival excess-inconsistent gingival margin | 68 | 45 | 66% | 23 | 34% | 0 | 0.0002 | 3.83 | 1.88 - 7.79 | F > M |
| Refractory periodontitis | 35 | 10 | 29% | 25 | 71% | 0 | 0.0005 | 0.16 | 0.06 - 0.45 | M > F |
| Gingivitis associated with pemphigus vulgaris | 34 | 25 | 74% | 9 | 26% | 0 | 0.0002 | 7.72 | 2.63 - 22.66 | F > M |
| **Dental caries** |  |  |  |  |  |  |  |  |  |  |
| Caries risk | 87845 | 46858 | 53% | 40553 | 46% | 434 | <0.0001 | 1.33 | 1.31 - 1.36 | F > M |
| Caries risk high | 65653 | 34577 | 53% | 30754 | 47% | 322 | <0.0001 | 1.26 | 1.24 - 1.29 | F > M |
| Primary caries (at DEJ) | 57537 | 30957 | 54% | 26192 | 46% | 388 | <0.0001 | 1.39 | 1.36 - 1.43 | F > M |
| Primary caries (<1/2 distance to the pulp) | 39646 | 21339 | 54% | 17919 | 45% | 388 | <0.0001 | 1.41 | 1.37 - 1.45 | F > M |
| Recurrent caries | 30655 | 17320 | 56% | 13206 | 43% | 128 | <0.0001 | 1.72 | 1.66 - 1.77 | F > M |
| Caries risk medium | 20416 | 11038 | 54% | 9292 | 46% | 86 | <0.0001 | 1.41 | 1.36 - 1.47 | F > M |
| Primary caries (>1/2 distance to the pulp) | 19362 | 10035 | 52% | 9170 | 47% | 157 | <0.0001 | 1.20 | 1.15 - 1.24 | F > M |
| Other dental caries | 14101 | 7303 | 52% | 6631 | 47% | 167 | <0.0001 | 1.21 | 1.16 - 1.27 | F > M |
| Recurrent caries (<1/2 distance to the pulp) | 12099 | 6850 | 57% | 5208 | 43% | 41 | <0.0001 | 1.73 | 1.64 - 1.82 | F > M |
| Dental caries, unspecified | 10733 | 5555 | 52% | 5044 | 47% | 134 | <0.0001 | 1.21 | 1.15 - 1.28 | F > M |
| Caries risk low | 9576 | 5255 | 55% | 4291 | 45% | 30 | <0.0001 | 1.50 | 1.42 - 1.59 | F > M |
| Root caries | 6447 | 3028 | 47% | 3390 | 53% | 29 | <0.0001 | 0.80 | 0.75 - 0.86 | M > F |
| Recurrent caries (>1/2 the distance to the pulp) | 5235 | 2895 | 55% | 2305 | 44% | 35 | <0.0001 | 1.57 | 1.46 - 1.70 | F > M |
| Incipient caries in enamel greater than 1/2 distance to DEJ | 4933 | 2693 | 55% | 2189 | 44% | 51 | <0.0001 | 1.51 | 1.39 - 1.63 | F > M |
| Recurrent caries (secondary caries)-(to the pulp) | 2374 | 1283 | 54% | 1073 | 45% | 18 | <0.0001 | 1.43 | 1.27 - 1.60 | F > M |
| Recurrent caries (secondary caries)-enamel | 1958 | 1183 | 60% | 772 | 39% | 3 | <0.0001 | 2.35 | 2.06 - 2.67 | F > M |
| Caries risk extreme | 1869 | 1006 | 54% | 858 | 46% | 5 | <0.0001 | 1.37 | 1.21 - 1.56 | F > M |
| Incipient caries in enamel unknown depth-NOS | 681 | 376 | 55% | 304 | 45% | 1 | 0.0001 | 1.53 | 1.23 - 1.89 | F > M |
| **Malignant tumor of oral cavity** |  |  |  |  |  |  |  |  |  |  |
| Malignant neoplasm | 616 | 258 | 42% | 358 | 58% | 0 | <0.0001 | 0.52 | 0.41 - 0.65 | M > F |
| **Diseases of the oral soft tissues, oral mucosa, tongue, and salivary glands** |  |  |  |  |  |  |  |  |  |  |
| Soft tissue calcification | 2141 | 1143 | 53% | 983 | 46% | 15 | <0.0001 | 1.35 | 1.19 - 1.52 | F > M |
| White-yellow lesion | 1791 | 1042 | 58% | 744 | 42% | 5 | <0.0001 | 1.96 | 1.71 - 2.24 | F > M |
| Red-blue lesion | 1009 | 684 | 68% | 323 | 32% | 2 | <0.0001 | 4.47 | 3.71 - 5.39 | F > M |
| Tongue swelling/pathology | 892 | 583 | 65% | 306 | 34% | 3 | <0.0001 | 3.61 | 2.97 - 4.39 | F > M |
| Ulcerative condition | 682 | 405 | 59% | 276 | 40% | 1 | <0.0001 | 2.15 | 1.73 - 2.67 | F > M |
| xerostomia | 577 | 409 | 71% | 168 | 29% | 0 | <0.0001 | 5.93 | 4.60 - 7.64 | F > M |
| Vesiculobullous condition | 263 | 158 | 60% | 104 | 40% | 1 | <0.0001 | 2.30 | 1.62 - 3.26 | F > M |
| Buccal mucosal swelling | 256 | 154 | 60% | 101 | 39% | 1 | <0.0001 | 2.32 | 1.63 - 3.30 | F > M |
| Oral/perioral pigmentation | 198 | 125 | 63% | 72 | 36% | 1 | <0.0001 | 3.00 | 1.99 - 4.51 | F > M |
| **Disorders of tooth Development and eruption** |  |  |  |  |  |  |  |  |  |  |
| Abnormalities of teeth | 67895 | 32882 | 48% | 30977 | 46% | 4036 | <0.0001 | 1.12 | 1.10 - 1.14 | F > M |
| Occlusion disorders | 16357 | 9052 | 55% | 6830 | 42% | 475 | <0.0001 | 1.73 | 1.65 - 1.81 | F > M |
| Eruption abnormalities | 8663 | 4575 | 53% | 3670 | 42% | 418 | <0.0001 | 1.52 | 1.43 - 1.62 | F > M |
| Developed/Acquired defects | 5454 | 3161 | 58% | 2282 | 42% | 11 | <0.0001 | 1.92 | 1.78 - 2.07 | F > M |
| Impacted tooth | 4429 | 2335 | 53% | 1713 | 39% | 381 | <0.0001 | 1.77 | 1.63 - 1.92 | F > M |
| Enamel defect | 2830 | 1739 | 61% | 1086 | 38% | 5 | <0.0001 | 2.56 | 2.30 - 2.85 | F > M |
| Dentin defect | 2537 | 1636 | 64% | 889 | 35% | 12 | <0.0001 | 3.37 | 3.00 - 3.78 | F > M |
| Enamel and dentin defect | 441 | 264 | 60% | 176 | 40% | 1 | <0.0001 | 2.25 | 1.72 - 2.94 | F > M |
| Size alteration | 438 | 248 | 57% | 187 | 43% | 3 | <0.0001 | 1.75 | 1.34 - 2.29 | F > M |
| Cementum defect | 88 | 61 | 69% | 26 | 30% | 1 | <0.0001 | 5.39 | 2.83 - 10.26 | F > M |
| **Other disorders of teeth, hard tissues of teeth, and supporting structures** |  |  |  |  |  |  |  |  |  |  |
| Loss of tooth structure | 77435 | 40573 | 52% | 36479 | 47% | 383 | <0.0001 | 1.24 | 1.22 - 1.27 | F > M |
| Sensitive dentin | 2498 | 1614 | 65% | 872 | 35% | 12 | <0.0001 | 3.40 | 3.03 - 3.82 | F > M |
| Temporomandibular disorders | 1725 | 1304 | 76% | 417 | 24% | 4 | <0.0001 | 9.72 | 8.31 - 11.35 | F > M |
| Abrasion of teeth | 1221 | 658 | 54% | 556 | 46% | 7 | <0.0001 | 1.40 | 1.19 - 1.64 | F > M |
| Abrasion of teeth due to dentifrice | 435 | 246 | 57% | 189 | 43% | 0 | 0.0001 | 1.69 | 1.30 - 2.22 | F > M |
| Primary occlusal trauma | 420 | 271 | 65% | 149 | 35% | 0 | <0.0001 | 3.31 | 2.49 - 4.39 | F > M |
| Secondary occlusal trauma | 374 | 232 | 62% | 142 | 38% | 0 | <0.0001 | 2.67 | 1.99 - 3.59 | F > M |
| **Others<sup>‡</sup>** |  |  |  |  |  |  |  |  |  |  |
| Harmful oral habits | 5863 | 3536 | 60% | 2311 | 39% | 16 | <0.0001 | 2.34 | 2.17 - 2.52 | F > M |
| Deposits on/staining of teeth | 2042 | 1134 | 56% | 897 | 44% | 11 | <0.0001 | 1.59 | 1.19 - 1.34 | F > M |
| Systemic disease manifestation in jaw | 367 | 219 | 60% | 148 | 40% | 0 | <0.0001 | 2.19 | 1.63 - 2.94 | F > M |
| Lip swelling | 335 | 198 | 59% | 133 | 40% | 4 | <0.0001 | 2.20 | 1.61 - 2.99 | F > M |

\* reported as shown in BigMouth (total N = 4,686,402)
<sup>‡</sup> oral and dental disease concepts constituting the sole entry within their corresponding ICD-10-CM category in BigMouth
<sup>◊</sup> Others includes number of participants with self-reported 'other gender' and 'gender not reported'
<sup>#</sup> P-values ≤ 0.002 indicate statistical significance after Bonferroni correction; P-values between 0.003 and 0.05 indicate a trend for significance
OR: female to male odds ratio; CI: confidence interval
F: female; M: male

Concepts showing significant statistical evidence of male bias were malignant neoplasm (p≦ 0.0001; female to male OR = 0.52), generalized severe aggressive periodontitis (p≦ 0.0001; female to male OR = 0.75), root caries (p≦ 0.0001; female to male OR = 0.80) (Table 2). Additional concepts showing significant sex bias in BigMouth data are shown in Table 2. Concepts in which a trend for significance (0.0002 ≦ p ≦ 0.05) for sex bias or non-significant results were observed are shown in Appendix Table 2.

### Concepts showing sex differences in both All of Us and BigMouth

Among the concepts investigated in *All of Us* and BigMouth, complete term overlap was noted for 20 concepts, with 8 concepts showing discordant sex bias, namely ‘pulp degeneration/necrosis’, ‘chronic apical abscess’, ‘generalized moderate periodontitis’, ‘localized chronic periodontitis’, ‘dental caries penetrating into pulp’, ‘developmental abnormality of tooth size/form’, ‘attrition of teeth’, and ‘abrasion of teeth’ (Table 3).

**Table 3.** DOC concepts overlapping in All of Us and BigMouth showing sex bias.

| DOC concept | All of Us |  |  | BigMouth |  |  |
| --- | --- | --- | --- | --- | --- | --- |
|  | Sex Bias | OR | 95% CI | Sex Bias | OR | 95% CI |
| *Diseases of pulp and periapical tissues* |  |  |  |  |  |  |
| Dental abscess | F > M | 2.78 | 1.77 - 4.37 | F > M | 1.35 | 1.21 - 1.52 |
| Reversible pulpitis | F > M | 2.68 | 2.11 - 3.40 | F > M | 1.59 | 1.33 - 1.90 |
| Asymptomatic apical periodontitis | F > M | 2.00 | 1.59 - 2.54 | F > M | 1.67 | 1.53 - 1.81 |
| Symptomatic apical periodontitis | F > M | 1.63 | 1.40 - 1.90 | F > M | 2.77 | 2.61 - 2.93 |
| Pulp degeneration/Pulp necrosis | **M > F** | **0.44** | **0.25 - 0.78** | **F > M** | **1.36** | **1.31 - 1.42** |
| Chronic apical abscess | **M > F** | **0.48** | **0.37 - 0.63** | **F > M** | **1.36** | **1.24 - 1.49** |
| *Gingival and periodontal diseases* |  |  |  |  |  |  |
| Generalized moderate periodontitis | **M > F** | **0.02** | **0.02 - 0.04** | **F > M** | **1.12** | **1.08 - 1.15** |
| Localized chronic periodontitis | **M > F** | **0.18** | **0.14 - 0.23** | **F > M** | **1.42** | **1.36 - 1.48** |
| Chronic periodontitis | F > M | 1.33 | 1.23 - 1.45 | F > M | 1.11 | 1.09 - 1.13 |
| *Dental caries* |  |  |  |  |  |  |
| At high risk for dental caries | F > M | 2.25 | 1.28 - 3.96 | F > M | 1.33 | 1.31 - 1.36 |
| Dental caries | F > M | 1.64 | 1.56 - 1.72 | F > M | 1.21 | 1.15 - 1.28 |
| Dental caries penetrating the pulp | **M > F** | **0.55** | **0.46 - 0.65** | **F > M** | **1.10** | **1.03 - 1.18** |
| *Malignant tumor of oral cavity* |  |  |  |  |  |  |
| Malignant tumor/neoplasm of oral cavity | M > F | 0.50 | 0.41 - 0.61 | M > F | 0.52 | 0.41 - 0.65 |
| *Diseases of the oral soft tissues, oral mucosa, tongue, and salivary glands* |  |  |  |  |  |  |
| Xerostomia | F > M | 4.96 | 4.43 - 5.55 | F > M | 4.25 | 3.87 - 4.67 |
| Ulcerative condition of the mouth | F > M | 4.25 | 3.87 - 4.67 | F > M | 2.15 | 1.73 - 2.67 |
| *Disorders of tooth developmental and eruption* |  |  |  |  |  |  |
| Disturbances in tooth eruption | F > M | 5.14 | 4.49 - 5.89 | F > M | 1.52 | 1.43 - 1.62 |
| Developmental abnormality of tooth size/ form | **M > F** | **0.44** | **0.25 - 0.78** | **F > M** | **1.75** | **1.34 - 2.29** |
| Tooth disorder/abnormalities | F > M | 1.98 | 1.90 - 2.06 | F > M | 1.12 | 1.10 - 1.14 |
| *Other disorders of teeth, hard tissues of teeth, and supporting structures* |  |  |  |  |  |  |
| Attrition of teeth | **M > F** | **0.35** | **0.27 - 0.44** | **F > M** | **1.09** | **1.03 - 1.16** |
| Abrasion of teeth | **M > F** | **0.29** | **0.22 - 0.39** | **F > M** | **1.40** | **1.19 - 1.64** |

Bold font denotes differences in sex bias among the results from the two databases.
OR, female-to-male odds ratio; CI, Confidence Interval

## Discussion

In the present study, we compared the frequencies of more than 200 DOC disease concepts among 4,501,648 females and males residing in the United States utilizing two large databases (*All of Us* and BigMouth) featuring electronic health record data. Among the DOC disease concepts included in the analysis, significant sex-specific differences were found for 70% and 39% of the concepts found in *All of Us* and BigMouth, respectively. Specifically, a female predominance was identified in 54% and 97% of these dental concepts in each database. Instances of discordant sex bias among concepts in each database were also observed, albeit minimal.

Among the seven general concept categories in the *All of Us* database, ‘disorders of tooth development and eruption’, ‘diseases of the oral soft tissues, oral mucosa, tongue, and salivary glands’, and ‘diseases of pulp and periapical tissues’ were marked by female bias, meanwhile ‘gingival and periodontal diseases’, ‘malignant tumors of the oral cavity’, ‘dental caries’, and ‘other’ disorders were marked by male bias. Periodontal disease is widely known to affect males more frequently than females. A systematic review (Shiau and Reynolds 2010) and two US population-based studies (Albandar 2002) have confirmed a significantly male predilection in periodontal disease, with effect sizes ranging from 7.33 to 16.85. Potential explanations for this well-established sex difference may be attributed to biological, immunological, and behavioral reasons. For instance, sexual dimorphism has been reported to account for a more diverse oral microbiome, with males presenting an increased abundance of *Prevotella intermedia* compared to females (Vieira et al. 2017). Males also tend to demonstrate a more robust intrinsic inflammatory reaction to both injuries and microbial pathogens compared to females (Shiau and Reynolds 2010). This is evidenced by an increased toll like receptor (TLR)-4 signaling, distinct cytokine production (Klein and Flanagan 2016), and greater CD8+T cell count (Abdullah et al. 2012) in males. Moreover, risk factors commonly associated with periodontal disease (e.g., obesity, cardiovascular disease, type II diabetes), are also known to exhibit a male bias (Meisel et al. 2014; Regensteiner et al. 2015; Kim et al. 2020). Lastly, a study by Lipsky et al. (2021) suggested that men are less inclined to prioritize their oral health therefore increasing the potential to higher instances of periodontal disease. In contrast, ‘gingival and periodontal diseases’ in BigMouth were marked by female bias. Interestingly, these reflected specific gingivitis conditions and periodontitis manifestations frequently associated with autoimmune conditions. The observed higher female predilection could potentially be attributed to hormonal effects during pregnancy or menstrual cycles, as well as a more robust innate and adaptive immune response that makes females more vulnerable to autoimmune and anti-inflammatory conditions (Klein and Flanagan 2016).

Disorders of teeth, hard tissues of teeth, and supporting structures were also more prevalent in males from *All of Us*. Many of these included loss of teeth due to periodontal disease, which may result from a male bias for periodontal disease. Nevertheless, BigMouth data revealed a higher prevalence of these concepts in females. A possible explanation for such discrepancy is that the terminology for the dental concepts identified within the two databases examined were not completely standardized, and BigMouth often contained additional subtypes of each concept compared to *All of Us*. There might also be an inherent bias in BigMouth as the participants included mostly females.

Dental caries concepts exhibited a female bias in both databases, with females presenting between 1.3-to 1.64-fold higher risk of overall dental caries than males. When considering additional dental caries subtypes in *All of Us*, however, a marked male bias was observed for 7/9 concepts including carious exposure of pulp, dental caries on pit and fissure (in enamel, into dentin, and into pulp). In BigMouth, of the 18 dental caries-associated concepts, only root caries showed male bias. Female bias in dental caries has been supported by several narrative and systematic reviews (Lukacs and Largaespada 2006; Ferraro and Vieira 2010; Teshome et al. 2021). Moreover, an earlier tooth eruption in females, states of pregnancy, among others, have been suggested as potential reasons for a higher female prevalence for dental caries (Lukacs 2006). Additional studies support a similar prevalence of dental caries among the sexes (Shaffer et al. 2015; Skoskiewicz-Malinowska et al. 2021).

With the exception of ‘chronic apical abscess’ and ‘pulp and periapical disease’ that showed a male bias in All of Us, all concepts under ‘diseases of the pulp and periapical tissues’ category showed predominantly female bias in All of Us and BigMouth. Particularly, concepts associated with dental pain, such as toothache, symptomatic reversible pulpitis, and symptomatic apical periodontitis were at least 2 times more frequent in females than males. Similar prevalence rates and instances of either higher female or male predominance in pulp and periapical diseases have been documented in the literature (Berlink et al. 2015; Huumonen et al. 2017; Ferreira et al. 2022).

Similarly, both databases were concordant in identifying a female bias for all ‘diseases of oral soft tissues, tongue, and salivary glands’, with some concepts showing higher than 5-fold increased odds in females compared to males. Considering that many of these conditions have an autoimmune etiology, a higher female predisposition was expected (Klein and Flanagan 2016). Disorders of tooth development and eruption as well as dentofacial anomalies were also significantly more prevalent in females compared to males, similarly to previous reports (Letra et al. 2022).

Males displayed a significantly higher risk of oral malignancies compared to females in both databases. This has been consistently reported across various studies worldwide, where males exhibit between 1-to 5-fold increased risk compared to females (Ward et al. 2019; Park et al. 2022). Such higher male predisposition to cancer persists even after controlling for behavioral risk factors, including smoking and alcohol consumption (Park et al. 2022). This phenomenon has been attributed to the potential association between androgens and cancer development, whereas estrogens have been proposed to exert a protective effect (Park et al. 2022).

Increasing evidence has supported genetic predisposition as a contributor to sex-based biological differences in oral and dental diseases (Sangalli et al. 2023). In this context, female bias towards caries risk has been attributed to genetic variation in genes located on the X chromosome, which are implicated in amelogenesis, dietary preferences, oral microbiome composition, and salivary flow (Ferraro and Vieira 2010). Similarly, studies have reported that genetic polymorphisms may result in an altered host response to microbial challenges in the dental pulp therefore modulating caries progression and/or predisposition to pulp and periapical disease (Menezes-Silva et al. 2012; Messing et al. 2019; Petty et al. 2023). More Notably, sex-specific associations with polymorphisms in genes involved in inflammation, wound healing, bone remodeling, and apoptosis, have been reported for periapical disease (de Souza et al. 2019; Petty et al. 2023). In animal models, sexual dimorphism in immunological responses to oral infectious diseases, including differences in neutrophil trafficking, osteoclast differentiation and formation in cases of both periodontal and periapical disease has been widely reported (Valerio and Kirkwood 2018). Sex differences have also been reported for the extent of activation of innate and adaptive immune cells in females, including higher numbers of macrophages, dendritic cells, antigen-presenting cells, interferon activity, lymphocyte cells, CD4+T cells, and cytotoxic T cells (Klein and Flanagan 2016). Taken together, these multiple lines of evidence strongly support a role for sex as a biological variable underlying the pathogenesis of pulpal and periapical diseases, although non-biological factors may also play a role.

This study has numerous strengths but also has some limitations. Among the strengths is the large sample size surpassing previous studies and greatly enhancing the generalizability of our findings, as well as the large number and wide spectrum of DOC concepts investigated. Study limitations include inherent differences among the databases as each aggregates data from different sources and may contribute to occasional discrepant findings. For instance, *All of Us* contains medical and dental record data from individuals across the US who agree to share their data, whereas BigMouth data is directly imported from dental electronic health records from eleven participating dental schools. Hence, a lack of complete agreement between the participant community and concept terminologies in *All of Us* and BigMouth databases was expected. *All of Us* provided broader group categories, whereas BigMouth featured more concepts and very specific subcategories based on dental diagnostic terminology. Importantly, dental data from *All of Us* is more likely to be resultant of self-report or within electronic medical records whereas BigMouth data is retrieved from electronic dental records reflecting actual patient examinations. Among the main discordant findings of sex bias in the two databases were ‘pulp degeneration/necrosis’, ‘chronic apical abscess’, ‘generalized moderate and localized chronic periodontitis’, ‘dental caries penetrating the pulp’, ‘developmental anomaly of tooth size/form’, and ‘tooth abrasion’ and ‘attrition’. While these differences may have precluded more accurate comparisons between the databases, the overall similarities in the findings of female or male bias across many of the concepts investigated in the two databases are invaluable in identifying consistent pattern of sex disparities in oral and dental diseases and conditions on a large scale.

## Conclusions

Evidence supporting sex-specific differences DOC diseases and conditions is still scarce. This study corroborates previous reports suggesting sexual dimorphism in periodontal disease, dental caries, and malignant tumors of the oral cavity meanwhile providing new evidence for sexual dimorphism in pulpal and periapical disease, disorders of oral soft tissues and tooth development, and other DOC conditions. These findings provide valuable insight as they may be used to inform future studies aimed at uncovering the biological and non-biological factors that underlie sex-specific differences in disease, and towards clinical translation with sex-specific strategies to improve oral health of females and males. Further studies considering socio-economic and behavioral factors may also provide valuable insight into the role of sex differences in DOC diseases and conditions.

## Data Availability

All data produced in the present work are contained in the manuscript.

## Acknowledgements

We gratefully acknowledge *All of Us* participants for their contributions, without whom this research would not have been possible. We also thank the National Institutes of Health’s *All of Us* Research Program for making available the participant data examined in this study. Thanks to Dominic Woods for technical assistance. Emma Fetchko was supported by the University of Pittsburgh School of Dental Medicine Dean’s Summer Research Scholarship.

## Authors’ contributions

EF contributed to data acquisition and analysis and drafted the manuscript; LS contributed to data interpretation and drafted the manuscript; AL contributed to conception, data interpretation, and critically reviewed the manuscript.

## Reporting Guidelines

GATHER checklist for epidemiological reports on population estimates of health conditions was followed.

## APPENDIX

**Appendix Table 1.** Additional dental, oral and craniofacial concepts investigated in All of Us.

| EHR concepts* | Participant count | Female count | Male count | Other gender | P-value<sup>#</sup> | Sex bias (trend) |
| --- | --- | --- | --- | --- | --- | --- |
| dental caries extending into dentin | 2160 | 1040 | 980 | 60 | 0.07 | none |
| broken tooth without complication | 960 | 480 | 420 | <20 | 0.006 | F>M |
| fractured dental restorative material | 860 | 380 | 420 | <20 | 0.05 | none |
| fractured dental restorative material with loss of material | 720 | 320 | 360 | <20 | 0.03 | M>F |
| necrosis of the pulp | 700 | 320 | 360 | <20 | 0.03 | M>F |
| oral phase dysphagia | 640 | 320 | 280 | <20 | 0.03 | F>M |
| leukoplakia of oral mucosa | 600 | 280 | 300 | <20 | 0.25 | none |
| neoplasm of uncertain behavior of lip, oral cavity and pharynx | 420 | 200 | 220 | <20 | 0.17 | none |
| irreversible pulpitis | 360 | 180 | 180 | <20 | 1 | none |
| laceration of oral cavity | 320 | 140 | 140 | <20 | 1 | none |
| neoplasm of uncertain behavior of oral cavity | 240 | 120 | 120 | <20 | 1 | none |
| erosion of teeth | 220 | 100 | 120 | <20 | 0.06 | none |
| disturbance of oral epithelium | 180 | 80 | 100 | <20 | 0.04 | M>F |
| at risk for dental caries | 140 | 80 | 60 | <20 | 0.02 | F>M |
| failure of osseointegration of dental implant | 120 | 60 | 60 | <20 | 1 | none |
| dislocation of tooth | 120 | 60 | 60 | <20 | 1 | none |
| broken tooth with complication | 120 | 80 | 60 | <20 | 0.009 | F>M |
| cellulitis of oral soft tissues | 120 | 60 | 60 | <20 | 1 | none |
| pulp degeneration | 100 | 40 | 60 | <20 | 0.005 | M>F |
| developmental abnormality of tooth size and form | 100 | 40 | 60 | <20 | 0.005 | M>F |
| oral ulceration due to radiation burn | 100 | 40 | 60 | <20 | 0.005 | M>F |
| primary malignant neoplasm of floor of mouth | 100 | 40 | 60 | <20 | 0.005 | M>F |
| at high risk for dental caries | 100 | 60 | 40 | <20 | 0.005 | F>M |
| tooth loss | 80 | 40 | 40 | <20 | 1 | none |
| biological failure of dental implant after osseointegration | 60 | 40 | 40 | <20 | 1 | none |
| fracture of dental implant | 60 | 40 | 40 | <20 | 1 | none |

\* EHR, electronic health records reported as shown in All of Us (total N = 254,700)
<sup>#</sup> Chi-square test; P-values ≤ 0.0002 indicate statistical significance after Bonferroni correction; P-values between 0.0003 and 0.05 indicate trend for significance F: female; M: male

**Appendix Table 2.**
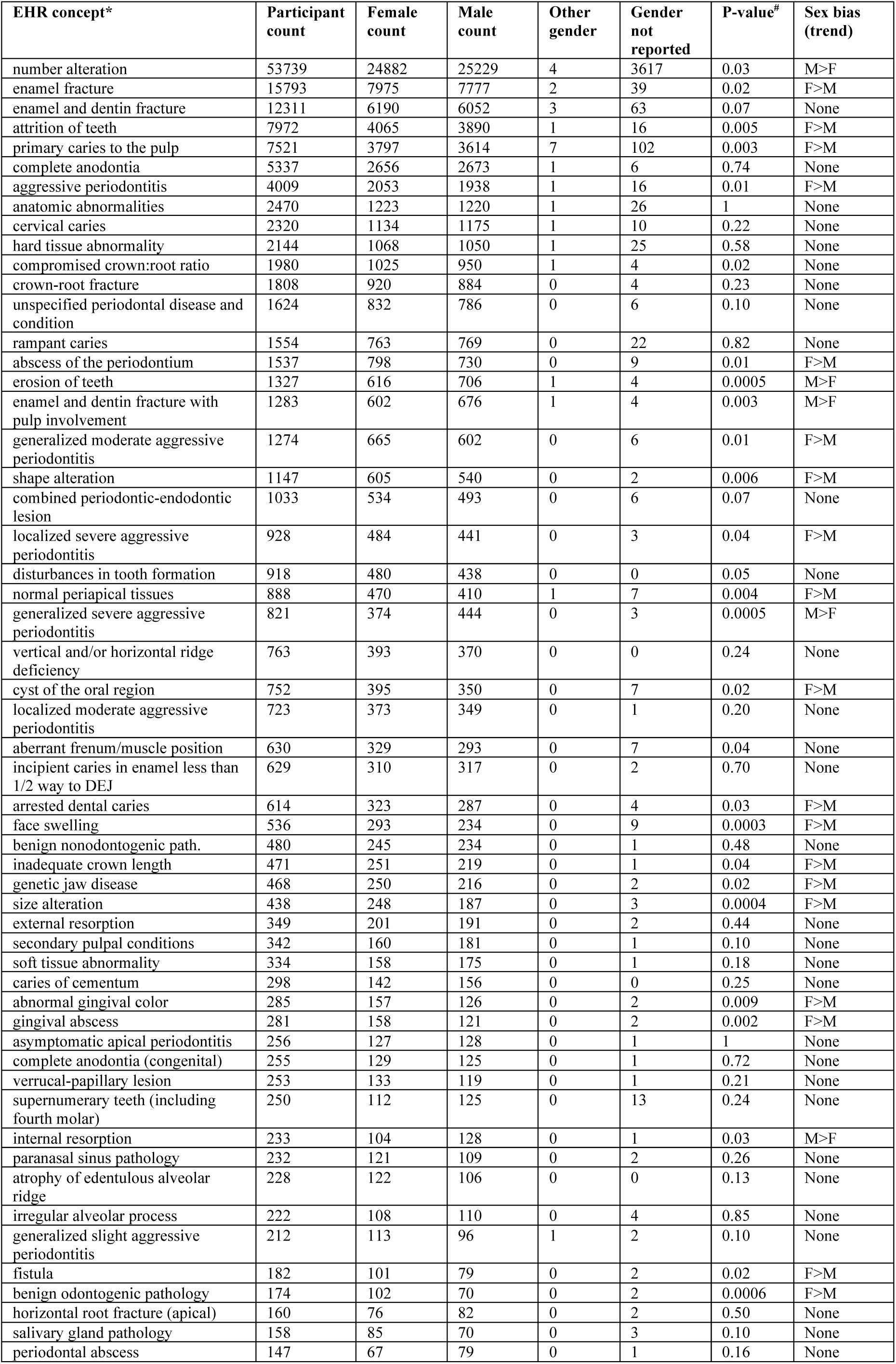
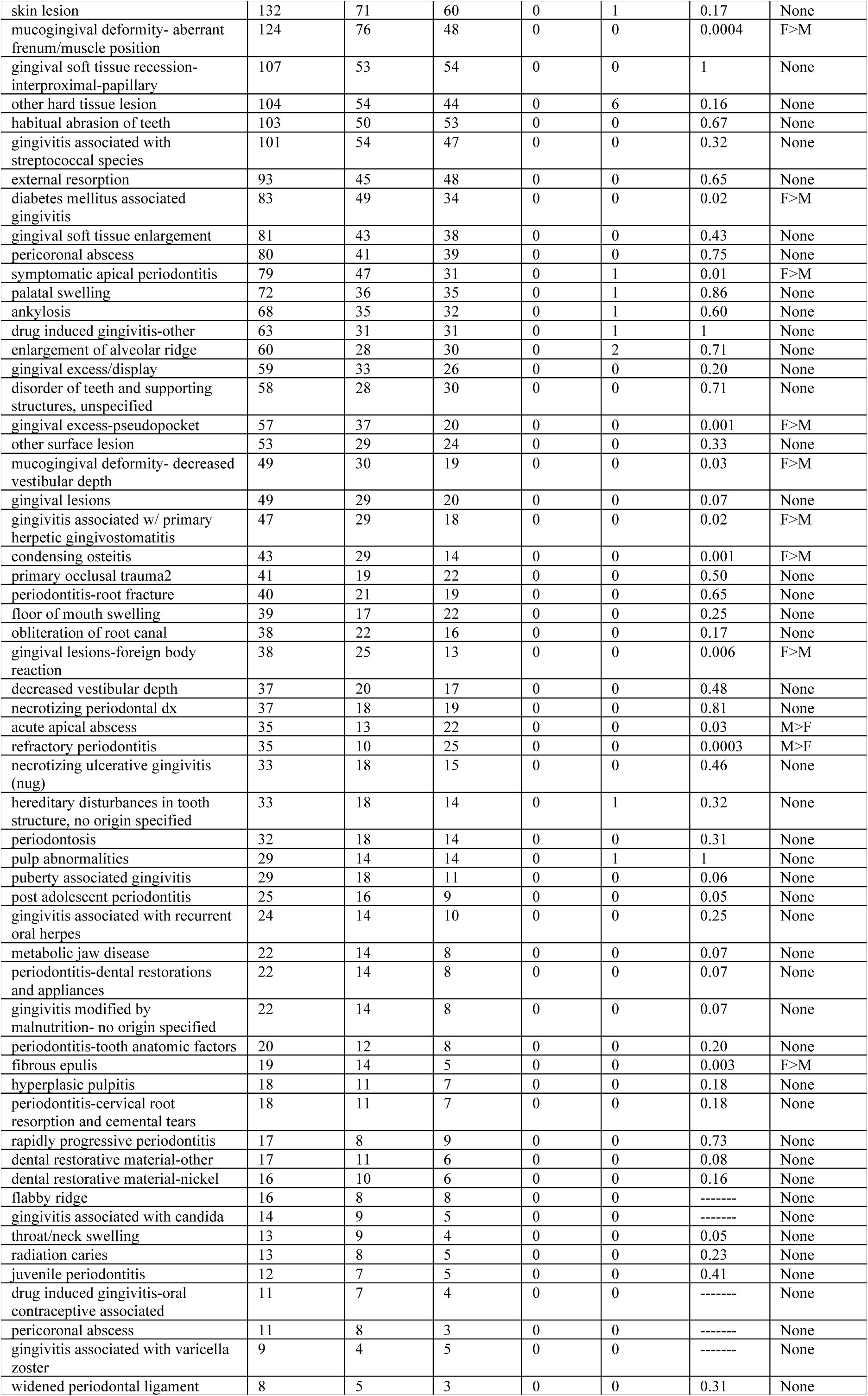
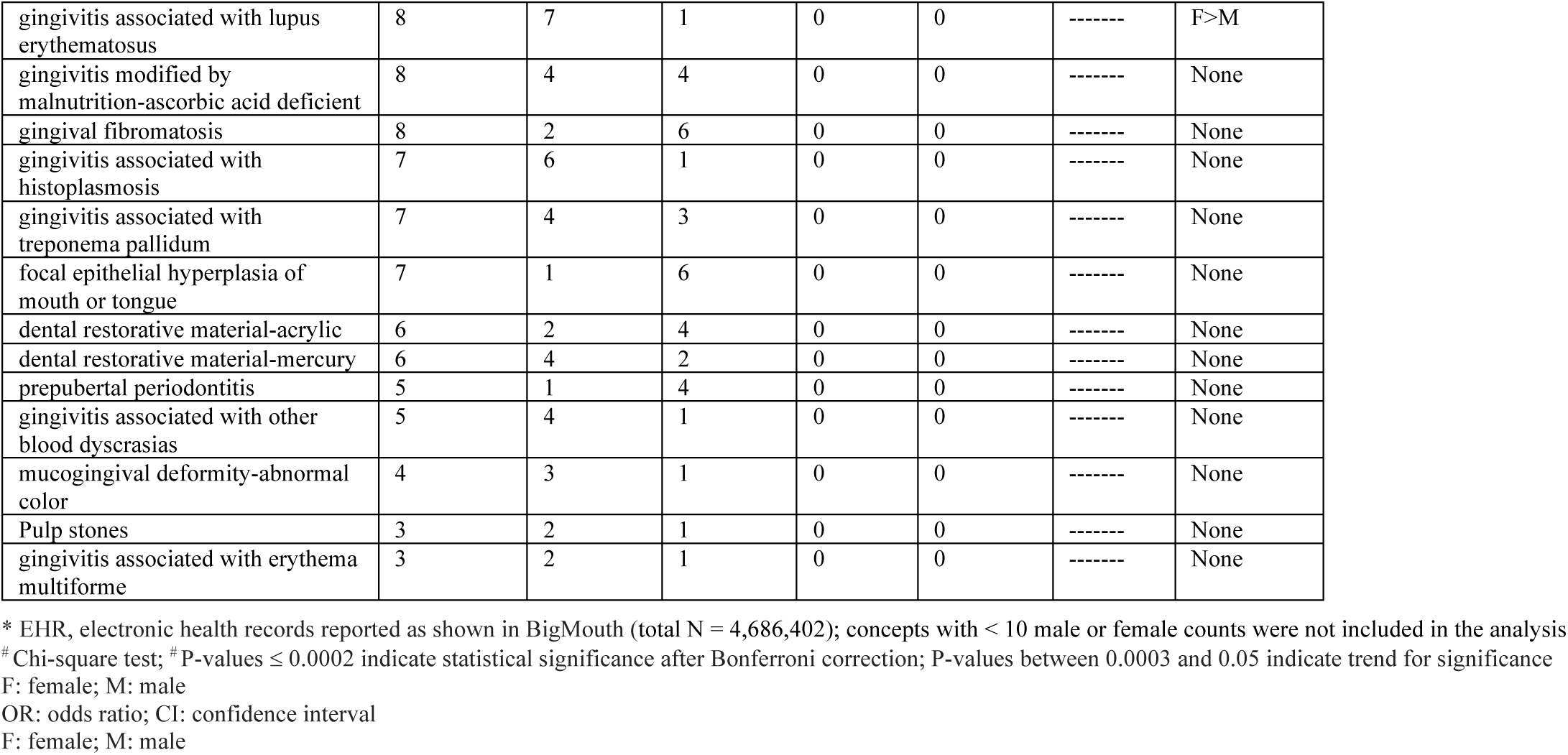
Additional dental, oral and craniofacial concepts investigated in BigMouth.

**Appendix Table 3.**
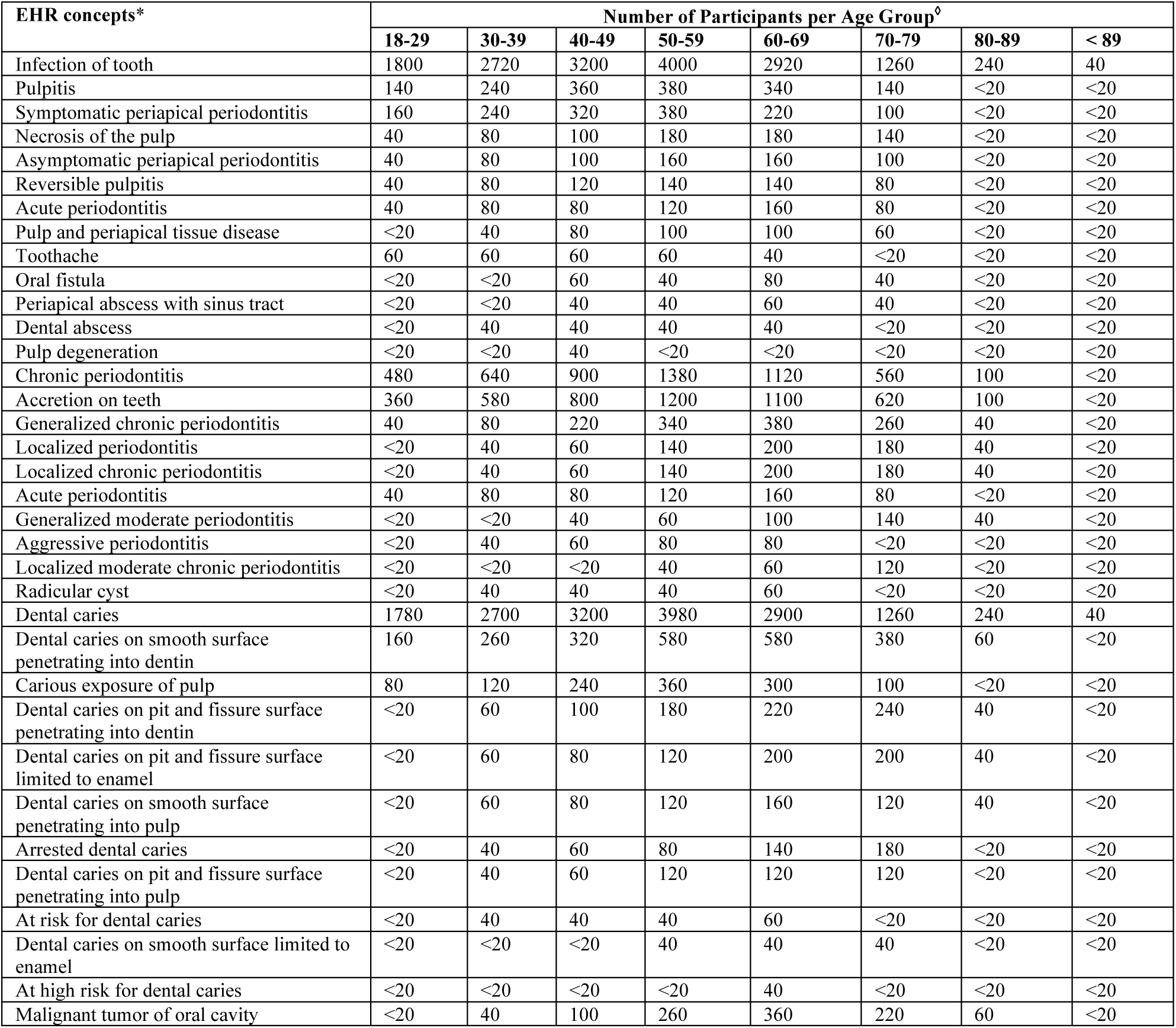
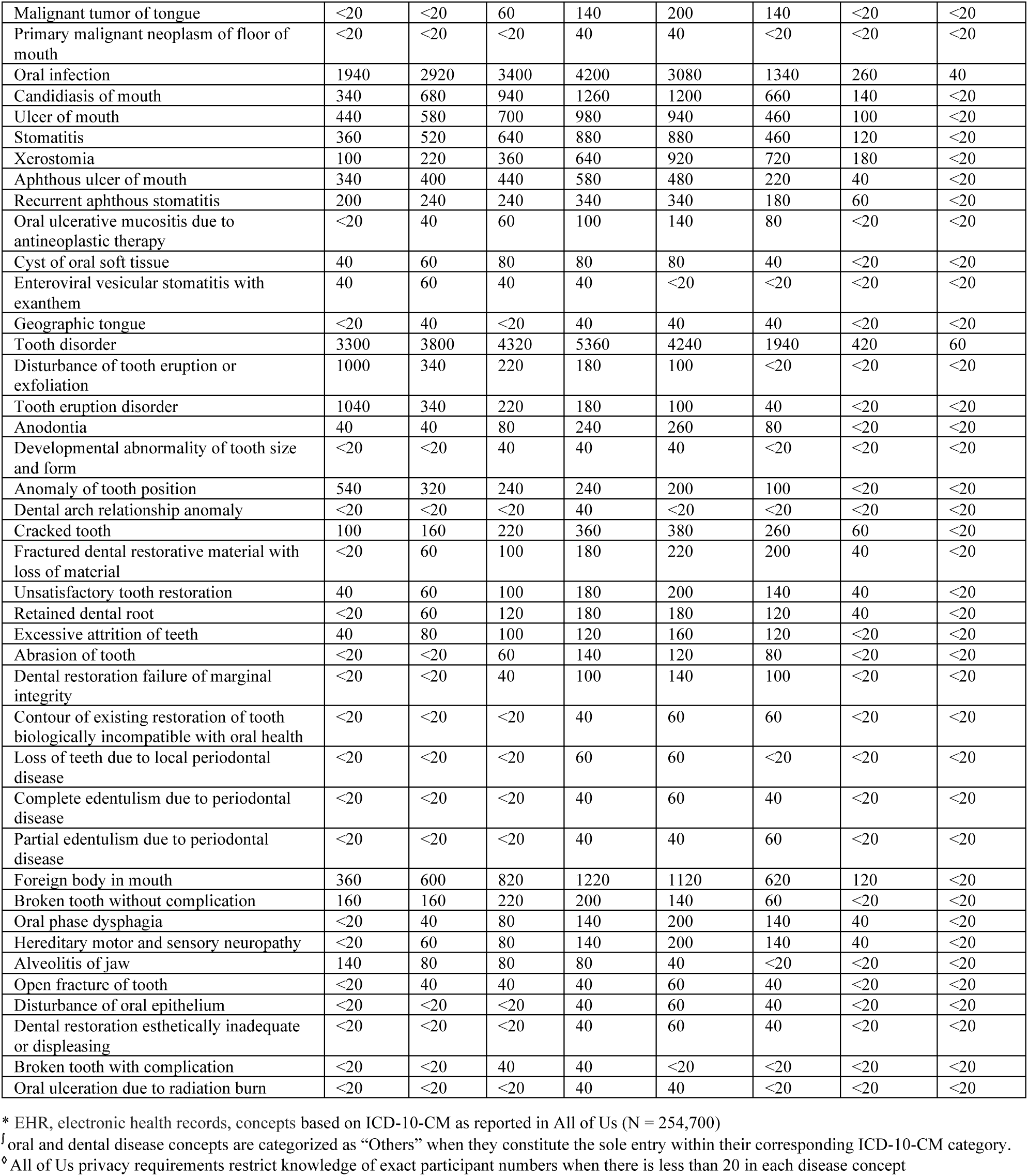
Number of participants per age group in All of Us according to each concept investigated.

| EHR concepts* | Number of Participants per Age Group<sup>o</sup> |  |  |  |  |  |  |  |
| --- | --- | --- | --- | --- | --- | --- | --- | --- |
|  | 18-29 | 30-39 | 40-49 | 50-59 | 60-69 | 70-79 | 80-89 | < 89 |
| Infection of tooth | 1800 | 2720 | 3200 | 4000 | 2920 | 1260 | 240 | 40 |
| Pulpitis | 140 | 240 | 360 | 380 | 340 | 140 | <20 | <20 |
| Symptomatic periapical periodontitis | 160 | 240 | 320 | 380 | 220 | 100 | <20 | <20 |
| Necrosis of the pulp | 40 | 80 | 100 | 180 | 180 | 140 | <20 | <20 |
| Asymptomatic periapical periodontitis | 40 | 80 | 100 | 160 | 160 | 100 | <20 | <20 |
| Reversible pulpitis | 40 | 80 | 120 | 140 | 140 | 80 | <20 | <20 |
| Acute periodontitis | 40 | 80 | 80 | 120 | 160 | 80 | <20 | <20 |
| Pulp and periapical tissue disease | <20 | 40 | 80 | 100 | 100 | 60 | <20 | <20 |
| Toothache | 60 | 60 | 60 | 60 | 40 | <20 | <20 | <20 |
| Oral fistula | <20 | <20 | 60 | 40 | 80 | 40 | <20 | <20 |
| Periapical abscess with sinus tract | <20 | <20 | 40 | 40 | 60 | 40 | <20 | <20 |
| Dental abscess | <20 | 40 | 40 | 40 | 40 | <20 | <20 | <20 |
| Pulp degeneration | <20 | <20 | 40 | <20 | <20 | <20 | <20 | <20 |
| Chronic periodontitis | 480 | 640 | 900 | 1380 | 1120 | 560 | 100 | <20 |
| Accretion on teeth | 360 | 580 | 800 | 1200 | 1100 | 620 | 100 | <20 |
| Generalized chronic periodontitis | 40 | 80 | 220 | 340 | 380 | 260 | 40 | <20 |
| Localized periodontitis | <20 | 40 | 60 | 140 | 200 | 180 | 40 | <20 |
| Localized chronic periodontitis | <20 | 40 | 60 | 140 | 200 | 180 | 40 | <20 |
| Acute periodontitis | 40 | 80 | 80 | 120 | 160 | 80 | <20 | <20 |
| Generalized moderate periodontitis | <20 | <20 | 40 | 60 | 100 | 140 | 40 | <20 |
| Aggressive periodontitis | <20 | 40 | 60 | 80 | 80 | <20 | <20 | <20 |
| Localized moderate chronic periodontitis | <20 | <20 | <20 | 40 | 60 | 120 | <20 | <20 |
| Radicular cyst | <20 | 40 | 40 | 40 | 60 | <20 | <20 | <20 |
| Dental caries | 1780 | 2700 | 3200 | 3980 | 2900 | 1260 | 240 | 40 |
| Dental caries on smooth surface penetrating into dentin | 160 | 260 | 320 | 580 | 580 | 380 | 60 | <20 |
| Carious exposure of pulp | 80 | 120 | 240 | 360 | 300 | 100 | <20 | <20 |
| Dental caries on pit and fissure surface penetrating into dentin | <20 | 60 | 100 | 180 | 220 | 240 | 40 | <20 |
| Dental caries on pit and fissure surface limited to enamel | <20 | 60 | 80 | 120 | 200 | 200 | 40 | <20 |
| Dental caries on smooth surface penetrating into pulp | <20 | 60 | 80 | 120 | 160 | 120 | 40 | <20 |
| Arrested dental caries | <20 | 40 | 60 | 80 | 140 | 180 | <20 | <20 |
| Dental caries on pit and fissure surface penetrating into pulp | <20 | 40 | 60 | 120 | 120 | 120 | <20 | <20 |
| At risk for dental caries | <20 | 40 | 40 | 40 | 60 | <20 | <20 | <20 |
| Dental caries on smooth surface limited to enamel | <20 | <20 | <20 | 40 | 40 | 40 | <20 | <20 |
| At high risk for dental caries | <20 | <20 | <20 | <20 | 40 | <20 | <20 | <20 |
| Malignant tumor of oral cavity | <20 | 40 | 100 | 260 | 360 | 220 | 60 | <20 |
| Malignant tumor of tongue | <20 | <20 | 60 | 140 | 200 | 140 | <20 | <20 |
| Primary malignant neoplasm of floor of mouth | <20 | <20 | <20 | 40 | 40 | <20 | <20 | <20 |
| Oral infection | 1940 | 2920 | 3400 | 4200 | 3080 | 1340 | 260 | 40 |
| Candidiasis of mouth | 340 | 680 | 940 | 1260 | 1200 | 660 | 140 | <20 |
| Ulcer of mouth | 440 | 580 | 700 | 980 | 940 | 460 | 100 | <20 |
| Stomatitis | 360 | 520 | 640 | 880 | 880 | 460 | 120 | <20 |
| Xerostomia | 100 | 220 | 360 | 640 | 920 | 720 | 180 | <20 |
| Aphthous ulcer of mouth | 340 | 400 | 440 | 580 | 480 | 220 | 40 | <20 |
| Recurrent aphthous stomatitis | 200 | 240 | 240 | 340 | 340 | 180 | 60 | <20 |
| Oral ulcerative mucositis due to antineoplastic therapy | <20 | 40 | 60 | 100 | 140 | 80 | <20 | <20 |
| Cyst of oral soft tissue | 40 | 60 | 80 | 80 | 80 | 40 | <20 | <20 |
| Enteroviral vesicular stomatitis with exanthem | 40 | 60 | 40 | 40 | <20 | <20 | <20 | <20 |
| Geographic tongue | <20 | 40 | <20 | 40 | 40 | 40 | <20 | <20 |
| Tooth disorder | 3300 | 3800 | 4320 | 5360 | 4240 | 1940 | 420 | 60 |
| Disturbance of tooth eruption or exfoliation | 1000 | 340 | 220 | 180 | 100 | <20 | <20 | <20 |
| Tooth eruption disorder | 1040 | 340 | 220 | 180 | 100 | 40 | <20 | <20 |
| Anodontia | 40 | 40 | 80 | 240 | 260 | 80 | <20 | <20 |
| Developmental abnormality of tooth size and form | <20 | <20 | 40 | 40 | 40 | <20 | <20 | <20 |
| Anomaly of tooth position | 540 | 320 | 240 | 240 | 200 | 100 | <20 | <20 |
| Dental arch relationship anomaly | <20 | <20 | <20 | 40 | <20 | <20 | <20 | <20 |
| Cracked tooth | 100 | 160 | 220 | 360 | 380 | 260 | 60 | <20 |
| Fractured dental restorative material with loss of material | <20 | 60 | 100 | 180 | 220 | 200 | 40 | <20 |
| Unsatisfactory tooth restoration | 40 | 60 | 100 | 180 | 200 | 140 | 40 | <20 |
| Retained dental root | <20 | 60 | 120 | 180 | 180 | 120 | 40 | <20 |
| Excessive attrition of teeth | 40 | 80 | 100 | 120 | 160 | 120 | <20 | <20 |
| Abrasion of tooth | <20 | <20 | 60 | 140 | 120 | 80 | <20 | <20 |
| Dental restoration failure of marginal integrity | <20 | <20 | 40 | 100 | 140 | 100 | <20 | <20 |
| Contour of existing restoration of tooth biologically incompatible with oral health | <20 | <20 | <20 | 40 | 60 | 60 | <20 | <20 |
| Loss of teeth due to local periodontal disease | <20 | <20 | <20 | 60 | 60 | <20 | <20 | <20 |
| Complete edentulism due to periodontal disease | <20 | <20 | <20 | 40 | 60 | 40 | <20 | <20 |
| Partial edentulism due to periodontal disease | <20 | <20 | <20 | 40 | 40 | 60 | <20 | <20 |
| Foreign body in mouth | 360 | 600 | 820 | 1220 | 1120 | 620 | 120 | <20 |
| Broken tooth without complication | 160 | 160 | 220 | 200 | 140 | 60 | <20 | <20 |
| Oral phase dysphagia | <20 | 40 | 80 | 140 | 200 | 140 | 40 | <20 |
| Hereditary motor and sensory neuropathy | <20 | 60 | 80 | 140 | 200 | 140 | 40 | <20 |
| Alveolitis of jaw | 140 | 80 | 80 | 80 | 40 | <20 | <20 | <20 |
| Open fracture of tooth | <20 | 40 | 40 | 40 | 60 | 40 | <20 | <20 |
| Disturbance of oral epithelium | <20 | <20 | <20 | 40 | 60 | 40 | <20 | <20 |
| Dental restoration esthetically inadequate or displeasing | <20 | <20 | <20 | 40 | 60 | 40 | <20 | <20 |
| Broken tooth with complication | <20 | <20 | 40 | 40 | <20 | <20 | <20 | <20 |
| Oral ulceration due to radiation burn | <20 | <20 | <20 | 40 | 40 | <20 | <20 | <20 |

\* EHR, electronic health records, concepts based on ICD-10-CM as reported in All of Us (N = 254,700)
<sup>f</sup> Oral and dental disease concepts are categorized as “Others” when they constitute the sole entry within their corresponding ICD-10-CM category.
<sup>g</sup> All of Us privacy requirements restrict knowledge of exact participant numbers when there is less than 20 in each disease concept

